# Social and health conditions that drive the need, demand, utilisation and expenditure on social care in the United Kingdom: a protocol for a systematic review

**DOI:** 10.1101/2024.08.19.24312217

**Authors:** Ben Amies-Cull, Sasha Shepperd, Nia Roberts, Anne Mason, Laura Bojke, Paul Clarkson, Anna Mae Scott

## Abstract

**Background:** Social care is the personal and practical assistance provided to individuals in their daily activities, personalised around their circumstances to promote wellbeing. It is provided by formal and informal carers, with formal care supported by considerable public resources through the Adult Social Care function of local authorities. While it is a matter of great public and policy concern that the system better meet people’s needs and that public finances are not unsustainably strained, some key drivers of adult social care need, such as the relationships between age, social determinants, environmental conditions, and health status are not well understood at the system level. This is a protocol for a systematic review of the evidence to determine the health and social drivers that contribute to adults’ need and demand for and utilisation of social care in the UK, and how these interact.

**Methods:** We will include quantitative studies of any experimental, observational or simulation/modelling design with average participant age ≥60, that examine the relationships between health status and/or social conditions, and their impact on adult social care need, demand, utilisation and expenditure. Informal and formal domiciliary, residential and nursing care, professional social work and occupational therapy will be included.

Medline, CINAHL, EconLit, ASSIA Campbell Collaboration and grey literature will be searched. A single reviewer will screen titles/abstracts for eligibility, and two reviewers will independently screen the full-text of studies initially considered eligible. 15% of the included studies will be double-extracted, and remainder single extracted with an accuracy check.

Risk of bias will be assessed using Cochrane Risk of Bias 2 and ROBINS-I.

The findings will be grouped by health condition(s), the type of determinant, outcome and will be presented in an evidence gap map. If three or more comparable studies are identified, we will consider calculating the effect size. We will use GRADE to assess the evidence certainty.

**Discussion:** We will detail the evidence on the relationships (to include an indication of their contribution) between health status and social conditions with the need, demand, utilisation and expenditure on adult social care in the UK, informing further analyses in key evidence gaps.

## Background and rationale

Social care includes Personal Care and other practical assistance to individuals with their activities of daily living(1) that encompasses a broad range of activities, ranging from general assistance with tasks such as laundry, cleaning, meal preparation, social activities, shopping, and assistance with dressing and washing.(2). It can also include professional assessments and support delivered by social workers and occupational therapists as part of the Adult Social Care function of local authorities to promote people’s wellbeing in as personalised way as possible.(3) Social care may be provided by professional or informal caregivers such as family, friends or neighbours.(4) It is estimated that approximately 20,000 organisations and over 5 million informal caregivers provide care for older people in the UK.(1)

Reasons for requiring formal social care vary, but as noted in the *Care Act 2014* – the most recent substantive legislation reforming social care in the United Kingdom (UK) – these include: “age, illness, disability, pregnancy, childbirth, dependence on alcohol or drugs, or any other similar circumstances”.(5, 6) Age alone is not specifically an indication for social care but is commonly associated with need, with 65% of recipients in the UK over 65 years old.(7)

Whilst it is recognised by governments that social care provision ought to be fair and accessible,(2) how best to accomplish this is unclear given the need to balance the elasticity of demand and the availability of resources. As state support in England, Wales and Northern Ireland is means tested, an individual’s personal resources are an important driver of the State’s social care costs. Scotland has free personal care for adults of any age, which was introduced in 2002 for those aged 65 and older and rolled out to all ages by 2019. (8) In Wales, non-residential social care is means-tested and individuals with savings and assets under 24,000 GBP are fully funded, whilst those with assets above that amount pay charges on a sliding scale to a maximum of 100 GBP per week.(9) In Northern Ireland, social care is the remit of the Department of Health. Following a care needs assessment, a means assessment determines the level of individual’s contribution towards the care cost. In England, above the upper threshold of assets and savings of 23,250 GBP, an individual is responsible for the full cost of care; between 14,250-23,250 GBP the individual is responsible for costs on a sliding scale and below 14,250 GBP the individual is eligible for fully funded care.(10)

In England, the responsibility for adult social care rests with local councils, which conduct a formal needs assessment with any relevant carer or proxy. This is within a national framework of minimum eligibility criteria and means testing.(11) Those with assets and savings below 14,250 GBP are expected to contribute their income to the costs of care, and those between 14,250-23,250 GBP are expected to provide an additional contribution that is proportionate to that capital on a sliding scale. Above 23,250 GBP in assets and savings, people are ineligible for publicly funded social care.(4, 7, 12). The value of an individual’s own home is excluded from the means test if the individual is receiving care in their home or a dependent remains living in that home when they are receiving residential care.(13) In practice, it was estimated in 2015 that lifetime costs of social care were 20,200-27,000 GBP for men and 38,700-49,000 GBP for women (in 2011/12 prices).(14) From October 2025, the current upper threshold of 23,250 GBP will increase to 100,000 GBP in savings.(13, 15)

Although social care is means tested in the UK, it nevertheless carries considerable public cost which is expected to grow. Councils spent 16.8 billion GBP on adult social care in 2016/17,(1) and it is estimated that from 2018 to 2038 the total expenditure on social care will increase by 94% to 1.25% of Gross Domestic Product.(16) Some of the contributing factors to increasing need include: increasing life expectancy (e.g. the number of over-85s is expected to increase from 1.3 million to 2.1 million between 2014 and 2030), a forecast increase in the years lived in poor health than the years lived in good health, an increase in the numbers of younger people with disabilities who are living longer (1, 17, 18) and long-term conditions, such as dementia, that incur a greater reliance on social care as they progress.(19)

The social conditions that people live in strongly determine their health status.(20–24) The evidence on ‘social determinants’ relates to both the presence of certain conditions and resources that are associated with downstream health impacts, as well as the unequal distribution of those factors between social groups.(25) In practice these are inseparable from health inequities.(24, 26) Social conditions include factors such as the quality of education, employment, income security, material conditions, the social safety-net, chronic stress, social exclusion, addiction and physical infrastructure.(23, 24, 27) As health status is an important predictor of social care need, the social determinants of health are likely to have downstream impacts on the quantity and distribution of need for social care, and therefore implications for access in the context of broadly insufficient supply.

Outside the relationship between health status and social care need, social conditions may independently impact on the quantity and distribution of social care need, demand, utilisation and expenditure. For example, adult children may provide informal care to relatives, and in more deprived areas may lack the resources to fulfil this role. Other barriers to accessing social care might also have a socio-economic dimension, such as digital literacy and ability to navigate the complicated care system. Self-funders have been found to enter the social care system earlier on average and with lower care needs, than those with less personal resources who are funded by local authorities and receive care later and with potentially higher care needs.(28) Some living conditions also influence the receipt of certain forms of social care, for example insanitary or poor accommodation can make admission to care homes more likely if the older person lives alone without carer support.(29)

It is important to consider “the causes of the causes” of social conditions, though downstream factors such as education have been critiqued as a “mid-level” policy target”(30) as they arise due to broader economic, social and political factors that represent the true social determinants (31) and their clustering in certain communities is not a coincidence. Describing and addressing more upstream factors remains challenging, but various taxonomies have been developed to help structure upstream, mid-level and downstream factors to target potential interventions. For example, the Diderichsen Model describes how social determinants can be broadly categorised as the causes of: i) social stratification, ii) differential exposure, ii) differential vulnerability and iv) differential consequences of illness.(32)

This review will consider the impact of social conditions and health on care need, demand, utilisation and expenditure. We will exclude studies that examine the effects of social determinants on health alone, as this work has been covered elsewhere.(20, 21, 23, 24, 26, 27) We will consider fully upstream and downstream factors and any point of potential policy intervention.

To enable sustainable, evidence-based, but locally-specific planning and decision-making around social care, it is crucial to understand what drives the need, demand, utilisation and expenditure on social care in the UK and the extent that this is driven by social conditions and health status. Within the context of increasing constraints on local authority budgets, understanding the social conditions that determine need, demand, utilisation and expenditure can inform how to avoid the current and future arrangements for social care from widening inequalities and impacting adversely on health and welfare. Currently, nearly one-third of requests for local government funding are declined, and service user contributions to care costs are increasing – including from those who are eligible for local authority funded care.(33)

The goal of the review is to identify and assess

1. The social conditions (e.g., health status, inequalities, education level, demographic characteristics, barriers to access, means-assessed access to social care, area deprivation, level of education, etc.) that drive the need, demand and utilisation of and expenditure (as one measure of utilisation) on formal and informal care in the UK
2. The impact of the social conditions’ and interaction on the need, use, demand, utilisation and expenditure on formal and informal social care.
3. The type, volume and certainty of evidence for each social condition on formal and informal social care need, demand, utilisation and expenditure.

The findings of this review will be used to:

- map and describe the certainty of evidence for different determinants of the need, demand, utilisation of and expenditure on formal and informal care;
- inform the identification of datasets required to complete an analysis of their collective impacts on social care;
- inform data analyses that will assess the drivers of requests for support for social care funding from local authorities;
- model the impacts of social determinants on need, demand, utilisation and/ or expenditure on social care;
- identify gaps in the literature base and generate hypotheses for further research.

## Methods

### Protocol development and availability

This protocol topic was informed by consultation with community members of the University of Oxford’s Nuffield Department of Population Health/REAL Demand Unit’s Patient and Public Involvement and Engagement Panel and social care stakeholders in UK.

The review is reported in compliance with the PRISMA-P checklist(34) (see supplementary material), and the patient and public engagement elements for this work will be reported using the Guidance for Reporting Involvement of Patients and the Public – Short Form (GRIPP2-SF).(35)

The protocol will be publicly available via medRxiv.

### Study Eligibility Criteria

#### Participants

We will include studies with adults with an average reported (mean, median, etc.) age of ≥60 years old. Studies of children, young adults, and adults with average age <60 years old will be excluded.

#### Concept

Social care has been defined as: “the personal care and support required by some people because of needs arising from their age, illness, disability or other circumstances.”(1) For the purposes of the present review, ‘social care’ will be considered broadly, i.e., if the study authors explicitly identify a social condition as one that is relevant to the need, or demand for, or utilisation of, or expenditure on, formal or informal social care, it will be treated as such.

We will include studies of health/ illness driving social care need, demand, utilisation and expenditure. Other social conditions of interest will include (but are not limited to): education level, demographic characteristics, employment status, social grade, household size, housing composition, isolation, functional status, area deprivation, barriers to access, and means-assessed access to social care.

#### Context

We will include studies that include participants who are living in their communities, in nursing/residential aged care homes, and those who movie into nursing/ residential care from hospital. This will include long and short-term care and rehabilitation.

We will include studies that are conducted in one or more of: England, Scotland, Wales, Northern Ireland, or the UK as a whole. We will include studies that are international as long as the social conditions that drive need, demand, utilisation and expenditure in the UK (or one of its constitutive nations), are reported separately from those in other jurisdictions.

We will include studies which report on health conditions/status and social conditions that drive need, demand, utilisation and expenditure on social care at any level: UK-wide, country-wide, regional or local within the UK.

We will exclude studies that report on social conditions driving health status (i.e., health as an outcome), as we are only interested in the need, demand, utilisation and expenditure of social care.

#### Study types

We will include studies identified from systematic reviews and scoping reviews, but will exclude literature reviews and other types of reviews.

We will include primary studies with the following designs:

1. Randomised trials of any design (individual randomisation, cluster, factorial, etc.), for example randomised trials of a community intervention that reported social conditions and measured use of social care
2. Longitudinal observational studies of any design (including but not restricted to prospective cohort studies, case-control studies, interrupted time series)
3. Cross sectional studies (single time point)
4. Simulation or other modelling studies

Qualitative studies will be excluded. Mixed methods studies (i.e. those including both quantitative and qualitative elements) will be included only if conclusions of the quantitative components can be clearly separated from the qualitative aspects of the study.

We will include data analysis studies (e.g. of linked data sets), and modelling studies based on ‘real data’ involving human participants.

### Search strategy

#### Searching for the published literature (databases)

The validation set of articles was obtained from content experts (see Appendix 1), and used to generate the preliminary search strategy. The Word Frequency Analyser (https://www.sr-accelerator.com/#/wordfreq) and MeSH On Demand (https://meshb.nlm.nih.gov/MeSHonDemand) tools were run on those articles to identify preliminary search terms. The strategy was drafted in PubMed, and SearchRefinery (https://www.sr-accelerator.com/#/searchrefinery) was used to adjust the precision of the search. The draft search strategy was circulated amongst the study team and stakeholders. Once feedback was received, the search string strategy was tested against the validation set of articles and further refined in SearchRefinery (by AMS) (see Appendix 2). The search strategy was reviewed by a search expert (NR) and finalised. For the searches in Medline, CINAHL, EconLit, an adapted version of the NICE UK geographic filter will be used(36); for ASSIA, the database’s filter for UK, England, Wales, Scotland or Northern Ireland will be used. For Campbell search, no geographic filter will be used. The full search strategies for all databases are provided in Appendix 3.

The search strategy was translated from Medline into other search databases using the Polyglot Search Translator (https://www.sr-accelerator.com/#/polyglot).

The following databases will be searched:

1. Medline (via PubMed)
2. CINAHL (via Ovid)
3. EconLit (via Ovid)
4. Applied Social Science Index and Abstracts, ASSIA (via ProQuest)
5. The Campbell Collaboration (via www.campbellcollaboration.org)

We will restrict the searches to articles published between 1 January 2009 and the present, to ensure the applicability of the findings to the present social care system and policy context in the UK, as there were major changes to the funding of Social Care, the NHS and local authorities from 2010.(37)

Only studies in English will be included, although given the focus of the review on the UK, it is not anticipated this will result in the exclusion of any studies.

#### Searching for grey literature

We will search the following sources for additional includable studies:

1. The websites for the health departments at the UK level and individual country-level (England, Scotland, Wales and Northern Ireland)
2. The websites of the following organisations: Social Care Institute for Excellence and the National Institute for Health and Care Excellence National Institutes for Health Research, Nuffield Trust, King’s Fund, The Health Foundation
3. Overton Knowledge Database

#### Forward (cited by) and backward (citing) search on included published studies

We will conduct a forward and backward citation search on the included studies using the SpiderCite tool (https://sr-accelerator.com/#/spidercite).

### Study screening and selection

The titles and abstracts will be single screened for inclusion/exclusion, and the full text of studies assessed as eligible will be independently dual screened. Where discrepancies or uncertainties arise during the title-abstract and full-text screening process, these will be assessed for inclusion by three reviewers. Published records (i.e., those identified in database searches) without an abstract will be screened in title only. Screening will be conducted in Covidence.

### Data charting

The data extraction form will be created jointly by the authors, who will then iteratively pilot it on 3 included studies at random. To ensure consistency in data extraction, 15% of the included studies will be double-extracted by two individuals, independently. Single extraction will be conducted for the remainder (85%) of the included studies, with a second reviewer checking the accuracy of the extraction. Where discrepancies or uncertainties arise during data extraction, these will be resolved by consensus or by referring to an additional author.

Extracted (charted) data will include:

1. General study characteristics: reference, study location (e.g., England, Wales, N. Ireland, Scotland), study design, study duration
2. Participant/population characteristics: N by gender and age, criteria for inclusion (population group and subgroups considered in the study), type of social care considered
3. Concept/exposure: health and social conditions e.g., category – inequality, demographics, health status, education, income level, housing; and the level at which the social condition is considered (local, regional, etc.)
4. Outcome: the need, demand or use of and expenditure on all formal or informal care.
5. Context: setting of the study (e.g. in the community, nursing/aged care homes, etc.)

### Data synthesis

The approach to data synthesis for each social condition will be as follows:

For all social conditions identified in the review, the volume and type of existing evidence will be mapped.(38, 39) The gap map will be generated using EPPI-Mapper, or another suitable package. The output will be presented in the style illustrated below, with each row corresponding to a social condition grouped by category (e.g. health, economic, demographic, etc.), and columns corresponding to the need, demand, utilisation and expenditure categories. Each cell will contain the information about the type and volume of evidence available, represented via a combination of bubble colours (different colour for each type of evidence – e.g. blue for interrupted time series, orange for RCTs, etc.) and bubble sizes (corresponding to the volume of the evidence – i.e. the higher the number of studies identified, the larger the bubble). An example is provided below (figure 1).

**Figure 1:**
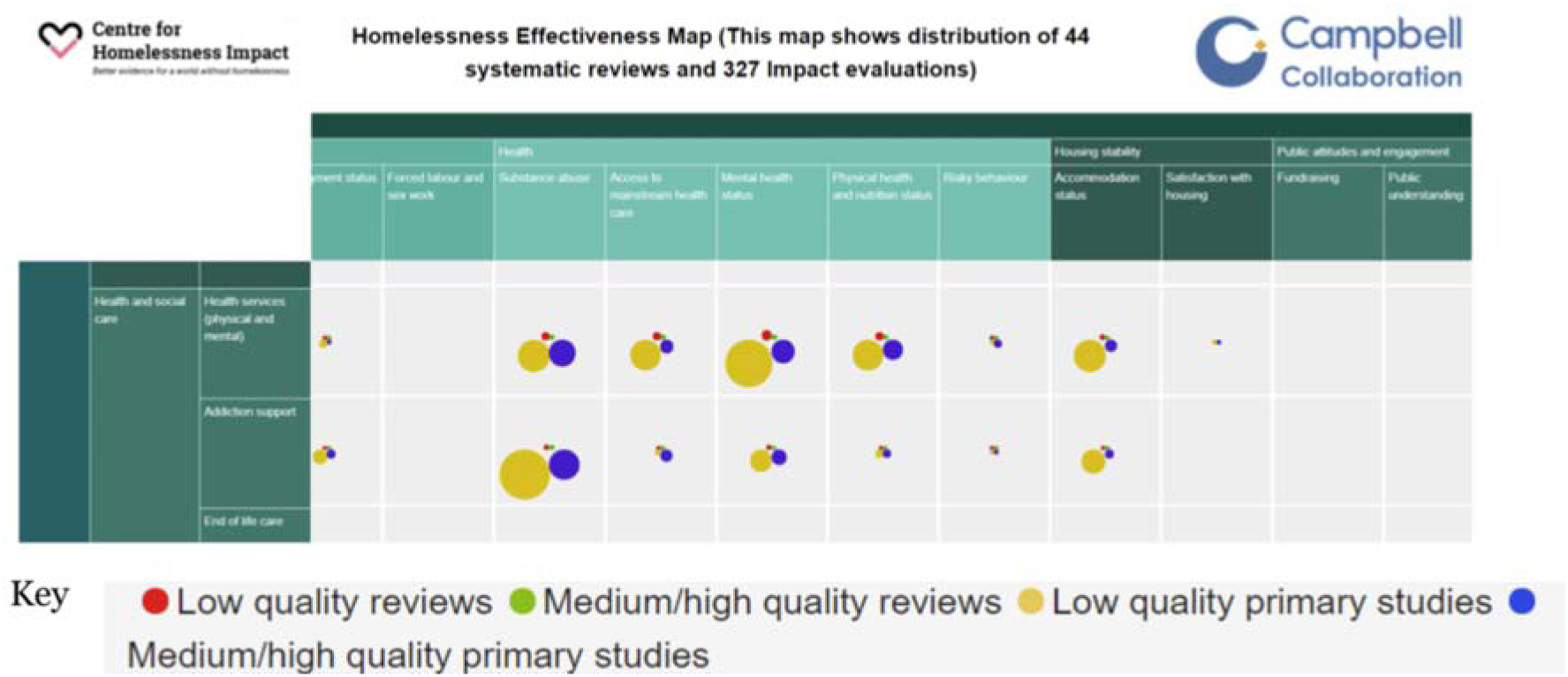
Example Evidence Gap Map output from the Campbell Collaboration(40)

**For social conditions for which ≥3 comparable studies with similar outcome data are identified**, Review Manager 5.4 will be used to calculate the effect size of each condition on the reported outcome (need, demand, utilisation of or expenditure on social care). For dichotomous outcomes, we will use risk ratios (where the number of individuals with an event is reported) or rate ratios (where the number of events and follow-up times are reported). For continuous outcomes, we will use mean difference (where the outcome is reported using the same scale across studies) or standardised mean difference (where outcome is reported using different scales across studies). Random effects models will be used due to expected heterogeneity of outcome measures.

If sufficient data are available for health conditions, we will subgroup outcomes by the specific health condition (e.g., diabetes, dementia, etc.) of the study population or combination of health conditions.

**Risk of bias** will be assessed for randomised trials by two review authors who will independently assess the risk of bias using Cochrane Risk of Bias Tool 2 for randomised trials and ROBINS-I for non-randomised studies.(41)

We will use GRADE to assess the certainty of the evidence for the contribution of each social condition. Two authors will independently apply the GRADE criteria; where judgements differ, consensus will be reached by discussion or by referring to a third author. The certainty will be ranked as very low, low, moderate or high. The certainty of the evidence for each health and social condition on outcome will be determined by considering eight GRADE factors, five of which may lead to rating down certainty (risk of bias, indirectness, inconsistency, imprecision, and publication bias) and three of which may result in rating up certainty (large effect, dose response, all plausible confounding and bias).(42)

## Data Availability

N/A - Review Protocol, no data available

## List of abbreviations

ASSIA: Applied Social Sciences Indexes and Abstracts
CINAHL: Cumulated Index to Nursing and Allied Health Literature
EPPI-mapper: Evidence for Policy and Practice Information-mapper
GRADE: Grading of Recommendations Assessment, Development and Evaluation
NHS: National Health Service
NICE: National Institute for Health and Care Excellence
PRISMA-P: Preferred Reporting Items for Systematic Review and Meta Analysis Protocols
RCT: Randomised Controlled Trial
REAL: Research and Economic Analysis for the Long-term
UK: United Kingdom

## Ethics and consent for participation

Not applicable. No ethics approval or consent was required.

## Consent for publication

Not applicable. No consent for publication was required.

## Availability of data and materials

Not applicable. No data were used in the manuscript. All methods materials are freely available as cited.

## Authors’ contributions

BAC conceived of the study, designed the methods and prepared the manuscript.

SS conceived of the study, designed the methods, prepared the manuscript and supervised the work.

NR designed the methods and reviewed the manuscript.

AM reviewed and provided feedback on the manuscript.

LB reviewed and provided feedback on the manuscript.

PC reviewed and provided feedback on the manuscript.

AMS conceived of the study, designed the methods, prepared the manuscript and supervised the work.

## Acknowledgements

We would like to thank the Research Support Services Specialist Centre for Social Care, for their advice and feedback. The REAL Demand Research Unit (University of Oxford) is funded by the Health Foundation’sCREAL Centre’s programme to develop evidence by conducting economic and health policy research to support better long-term decision making in health and social care.

## Appendix 1: Validation set of articles

6 references (reproduced below) were used as a validation set, to produce the search strategy and test its recall and precision.

Three of the references in the validation set would not meet the inclusion criteria of the present review due to being based in New Zealand rather than the UK (Wilkinson-Meyers 2013, Lay-Yee 2017) or pre-dating the date limits for the present review, which are 2009-present (Jette 1992). As these references were used for generating the search strategy and as part of a validation procedure, they are presented here for completeness.

1. Jette AM, Tennstedt SL, Branch LG. Stability of informal long-term care. Journal of aging and health. 1992;4(2):193-211.
2. Lay-Yee R, Pearson J, Davis P, von Randow M, Kerse N, Brown L. Changing the balance of social care for older people: simulating scenarios under demographic ageing in New Zealand. Health & social care in the community. 2017;25(3):962-74.
3. Lyu JY, Hu B, Wittenberg R, King D. The relationships between informal and formal social care for older people in England: A comparison before and after the Care Act 2014. Journal of aging & social policy. 2023:1-18.
4. MacLeod CA, Bu F, Rutherford AC, Phillips J, Woods R. Cognitive impairment negatively impacts allied health service uptake: Investigating the association between health and service use. SSM - population health. 2021;13:100720.
5. Subbe CP, Goulden N, Mawdsley K, Smith R. Anticipating care needs of patients after discharge from hospital: Frail and elderly patients without physiological abnormality on day of admission are more likely to require social services input. European journal of internal medicine. 2017;45:74-7.
6. Wilkinson-Meyers L, Brown P, McLean C, Kerse N. Met and unmet need for personal assistance among community-dwelling New Zealanders 75 years and over. Health & social care in the community. 2014;22(3):317-27.

## Appendix 2 – Search strategy testing (Search Refinery)

### Test of the “Driver” terms

- String tested: “predict*”[Title/Abstract] OR “determinant*”[Title/Abstract] OR “driver*”[Title/Abstract] OR “projecti*”[Title/Abstract] OR “promot*”[Title/Abstract]
- PMIDs for the validation set of 6: (28974330 or 33364299 or 10117873 or 24354894 or 27709717 or 37353920)
- The string finds all 6 of the relevant citations in the validation set

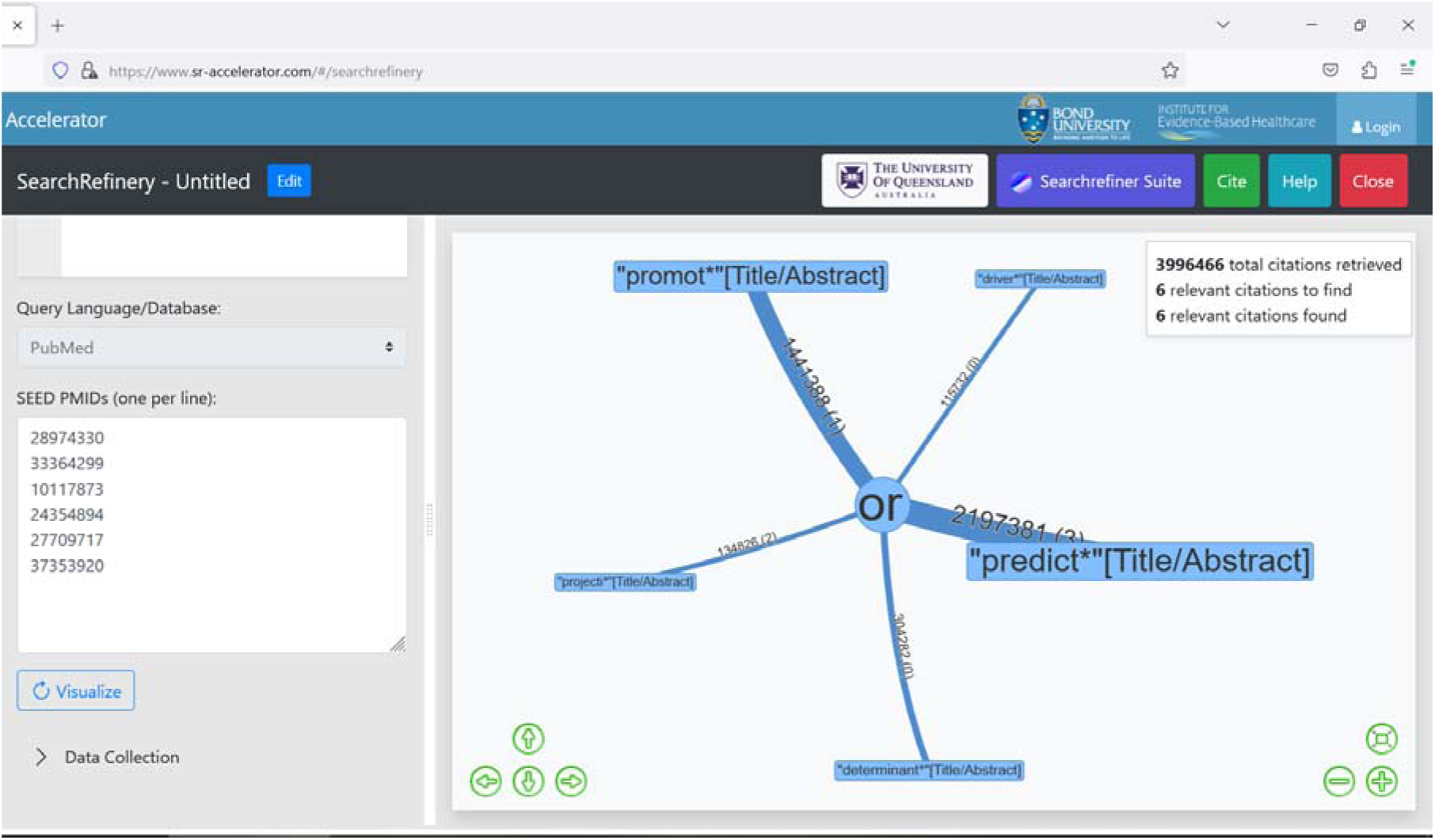

### Test of the “need / demand / utilisation / expenditure” terms

- String tested: “Health Services Needs and Demand”[MeSH Terms:noexp] OR “need*”[Title/Abstract] OR “demand*”[Title/Abstract] OR “using”[Title/Abstract] OR “utilis*”[Title/Abstract] OR “utiliz*”[Title/Abstract] OR “access*”[Title/Abstract] OR “Health Expenditures”[MeSH Terms:noexp] OR “resourc*”[Title/Abstract]
- PMIDs for the validation set of 6: (28974330 or 33364299 or 10117873 or 24354894 or 27709717 or 37353920)
- The string finds all 6 of the relevant citations in the validation set

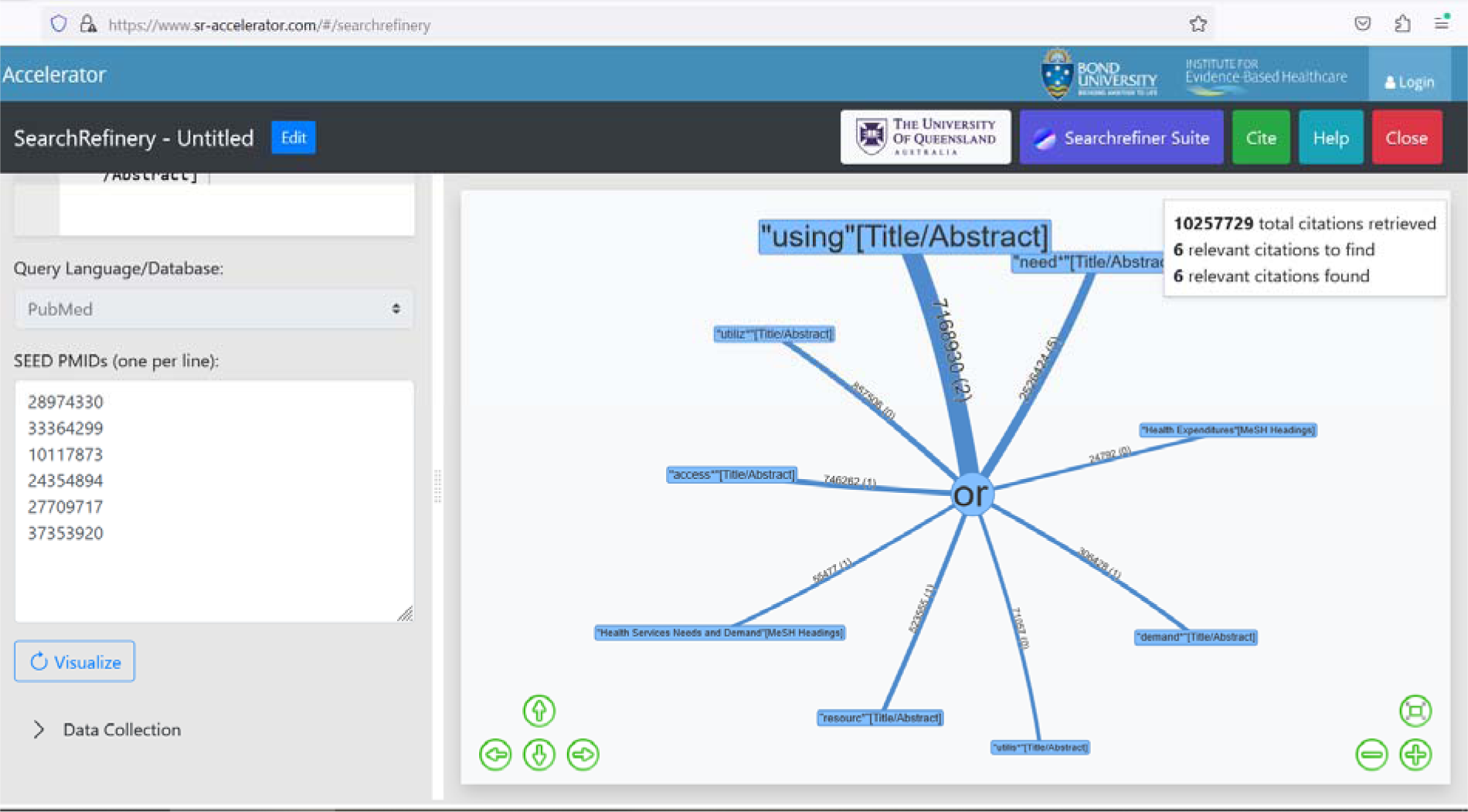

### Test of the “social care” terms

- String tested: “long term care”[MeSH Terms:noexp] OR “long term care”[Title/Abstract] OR “Social Work”[MeSH Terms:noexp] OR “social work*”[Title/Abstract] OR “formal care”[Title/Abstract] OR “informal care”[Title/Abstract] OR “Home Care Services”[MeSH Terms:noexp] OR “home care”[Title/Abstract] OR “homecare”[Title/Abstract] OR “communit*”[Title/Abstract] OR “Community Health Services”[MeSH Terms:noexp] OR “social service*”[Title/Abstract] OR “social care”[Title/Abstract]
- PMIDs for the validation set of 6: (28974330 or 33364299 or 10117873 or 24354894 or 27709717 or 37353920)
- The string finds all 6 of the relevant citations in the validation set

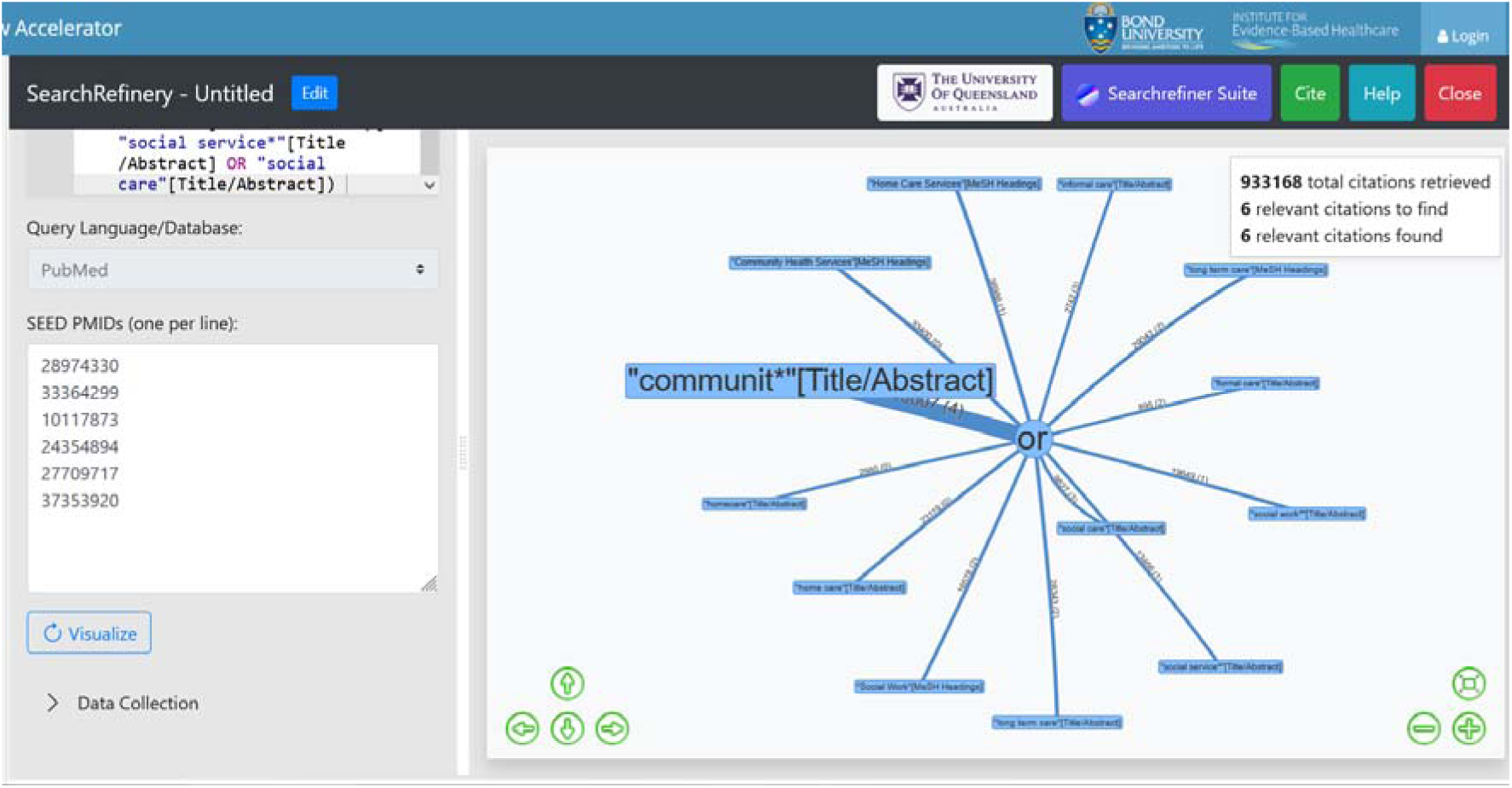

## Appendix 3: Search strategies

### Medline (via PubMed)

((“predict*”[Title/Abstract] OR “determinant*”[Title/Abstract] OR “driver*”[Title/Abstract] OR “projecti*”[Title/Abstract] OR “promot*”[Title/Abstract]))

AND

(“Health Services Needs and Demand”[MeSH Terms:noexp] OR “need*”[Title/Abstract] OR “demand*”[Title/Abstract] OR “using”[Title/Abstract] OR “utilis*”[Title/Abstract] OR “utiliz*”[Title/Abstract] OR “access*”[Title/Abstract] OR “Health Expenditures”[MeSH Terms:noexp] OR “resourc*”[Title/Abstract])

AND

(“long term care”[MeSH Terms:noexp] OR “long term care”[Title/Abstract] OR “Social Work”[MeSH Terms:noexp] OR “social work*”[Title/Abstract] OR “formal care”[Title/Abstract] OR “informal care”[Title/Abstract] OR “Home Care Services”[MeSH Terms:noexp] OR “home care”[Title/Abstract] OR “homecare”[Title/Abstract] OR “communit*”[Title/Abstract] OR “Community Health Services”[MeSH Terms:noexp] OR “social service*”[Title/Abstract] OR “social care”[Title/Abstract])

AND

(2009/01/01:2024/12/31[dp])

AND

((“United Kingdom”[MeSH Terms] OR (“national health service*”[Title/Abstract] OR “nhs”[Title/Abstract]) OR (“gb”[Title/Abstract] OR “g b”[Title/Abstract] OR “britain*”[Title/Abstract] OR (“british*”[Title/Abstract] NOT “british columbia”[Title/Abstract]) OR “uk”[Title/Abstract] OR “u k”[Title/Abstract] OR “united kingdom*”[Title/Abstract] OR (“england*”[Title/Abstract] NOT “new england”[Title/Abstract]) OR “northern ireland*”[Title/Abstract] OR “northern irish*”[Title/Abstract] OR “scotland*”[Title/Abstract] OR “scottish*”[Title/Abstract] OR ((“wales”[Title/Abstract] OR “south wales”[Title/Abstract]) NOT “new south wales”[Title/Abstract]) OR “welsh*”[Title/Abstract]) OR (“bath”[Title/Abstract] OR “bath’s”[Title/Abstract] OR ((“birmingham”[Title/Abstract] NOT “alabama*”[Title/Abstract]) OR (“birmingham’s”[Title/Abstract] NOT “alabama*”[Title/Abstract]) OR “bradford”[Title/Abstract] OR “bradford’s”[Title/Abstract] OR “brighton”[Title/Abstract] OR “brighton’s”[Title/Abstract] OR “bristol”[Title/Abstract] OR “bristol’s”[Title/Abstract] OR “carlisle*”[Title/Abstract] OR (“cambridge”[Title/Abstract] NOT (“massachusetts*”[Title/Abstract] OR “boston*”[Title/Abstract] OR “harvard*”[Title/Abstract])) OR (“canterbury”[Title/Abstract] NOT “zealand*”[Title/Abstract]) OR “chelmsford”[Title/Abstract] OR “chester”[Title/Abstract] OR “chester’s”[Title/Abstract] OR “chichester”[Title/Abstract] OR “coventry”[Title/Abstract] OR “coventry’s”[Title/Abstract] OR “derby”[Title/Abstract] OR “derby’s”[Title/Abstract] OR (“durham”[Title/Abstract] NOT (“carolina*”[Title/Abstract] OR “nc”[Title/Abstract])) OR (“durham’s”[Title/Abstract] NOT (“carolina*”[Title/Abstract] OR “nc”[Title/Abstract])) OR “ely”[Title/Abstract] OR “ely’s”[Title/Abstract] OR “exeter”[Title/Abstract] OR “gloucester”[Title/Abstract] OR “hereford”[Title/Abstract] OR “hull”[Title/Abstract] OR “hull’s”[Title/Abstract] OR “lancaster”[Title/Abstract] OR “lancaster’s”[Title/Abstract] OR “leeds*”[Title/Abstract] OR “leicester”[Title/Abstract] OR (“lincoln”[Title/Abstract] NOT “nebraska*”[Title/Abstract]) OR (“lincoln’s”[Title/Abstract] NOT “nebraska*”[Title/Abstract]) OR (“liverpool”[Title/Abstract] NOT (“new south wales*”[Title/Abstract] OR “nsw”[Title/Abstract])) OR (“liverpool’s”[Title/Abstract] NOT (“new south wales*”[Title/Abstract] OR “nsw”[Title/Abstract])) OR ((“london”[Title/Abstract] NOT (“ontario*”[Title/Abstract] OR “ont”[Title/Abstract] OR “toronto*”[Title/Abstract])) OR (“london’s”[Title/Abstract] NOT (“ontario*”[Title/Abstract] OR “ont”[Title/Abstract] OR “toronto*”[Title/Abstract])) OR “manchester”[Title/Abstract] OR “manchester’s”[Title/Abstract] OR (“newcastle”[Title/Abstract] NOT (“new south wales*”[Title/Abstract] OR “nsw”[Title/Abstract])) OR (“newcastle’s”[Title/Abstract] NOT (“new south wales*”[Title/Abstract] OR “nsw”[Title/Abstract])) OR “norwich”[Title/Abstract] OR “nottingham”[Title/Abstract] OR “nottingham’s”[Title/Abstract] OR “oxford”[Title/Abstract] OR “oxford’s”[Title/Abstract] OR “peterborough”[Title/Abstract] OR “plymouth”[Title/Abstract] OR “plymouth’s”[Title/Abstract] OR “portsmouth”[Title/Abstract] OR “preston”[Title/Abstract] OR “preston’s”[Title/Abstract] OR “ripon”[Title/Abstract] OR “salford”[Title/Abstract] OR “salisbury”[Title/Abstract] OR “salisbury’s”[Title/Abstract] OR “sheffield”[Title/Abstract] OR “sheffield’s”[Title/Abstract] OR “southampton”[Title/Abstract] OR “southampton’s”[Title/Abstract] OR “st albans”[Title/Abstract] OR “stoke”[Title/Abstract] OR “stoke’s”[Title/Abstract] OR “sunderland”[Title/Abstract] OR “sunderland’s”[Title/Abstract] OR “truro”[Title/Abstract] OR “wakefield”[Title/Abstract] OR “wakefield’s”[Title/Abstract] OR “wells”[Title/Abstract] OR “westminster”[Title/Abstract] OR “winchester”[Title/Abstract] OR “wolverhampton”[Title/Abstract] OR (“worcester”[Title/Abstract] NOT (“massachusetts*”[Title/Abstract] OR “boston*”[Title/Abstract] OR “harvard*”[Title/Abstract])) OR (“york”[Title/Abstract] NOT (“new york*”[Title/Abstract] OR “ny”[Title/Abstract] OR “ontario*”[Title/Abstract] OR “ont”[Title/Abstract] OR “toronto*”[Title/Abstract])) OR (“york’s”[Title/Abstract] NOT (“new york*”[Title/Abstract] OR “ny”[Title/Abstract] OR “ontario*”[Title/Abstract] OR “ont”[Title/Abstract] OR “toronto*”[Title/Abstract]))))) OR (“bangor”[Title/Abstract] OR “cardiff”[Title/Abstract] OR “newport”[Title/Abstract] OR “newport’s”[Title/Abstract] OR “st asaph”[Title/Abstract] OR “st davids”[Title/Abstract] OR “swansea”[Title/Abstract]) OR (“aberdeen”[Title/Abstract] OR “dundee”[Title/Abstract] OR “dundee’s”[Title/Abstract] OR “edinburgh”[Title/Abstract] OR “edinburgh’s”[Title/Abstract] OR “glasgow”[Title/Abstract] OR “glasgow’s”[Title/Abstract] OR “inverness”[Title/Abstract] OR (“perth”[Title/Abstract] NOT “australia*”[Title/Abstract]) OR (“perth’s”[Title/Abstract] NOT “australia*”[Title/Abstract]) OR “stirling”[Title/Abstract] OR “stirling’s”[Title/Abstract]) OR (“armagh”[Title/Abstract] OR “belfast”[Title/Abstract] OR “lisburn”[Title/Abstract] OR “londonderry”[Title/Abstract] OR “derry”[Title/Abstract] OR “derry’s”[Title/Abstract] OR “newry”[Title/Abstract])) NOT (“africa”[MeSH Terms] OR “americas”[MeSH Terms] OR “antarctic regions”[MeSH Terms] OR “arctic regions”[MeSH Terms] OR “asia”[MeSH Terms] OR “oceania”[MeSH Terms])) NOT (“United Kingdom”[MeSH Terms] OR “europe”[MeSH Terms:noexp])

### CINAHL (via Ebsco)

((TI predict* OR AB predict*) OR (TI determinant* OR AB determinant*) OR (TI driver* OR AB driver*) OR (TI projecti* OR AB projecti*) OR (TI promot* OR AB promot*))

AND

((MH “Health Services Needs and Demand”) OR (TI need* OR AB need*) OR (TI demand* OR AB demand*) OR (TI using OR AB using) OR (TI utilis* OR AB utilis*) OR (TI utiliz* OR AB utiliz*) OR (TI access* OR AB access*) OR (MH “Health Expenditures”) OR (TI resourc* OR AB resourc*))

AND

((MH “long term care”) OR (TI “long term care” OR AB “long term care”) OR (MH “Social Work”) OR (TI “social work*” OR AB “social work*”) OR (TI “formal care” OR AB “formal care”) OR (TI “community care” OR AB “community care”) OR (TI “informal care” OR AB “informal care”) OR (MH “Home Care Services”) OR (TI “home care” OR AB “home care”) OR (TI homecare OR AB homecare) OR (MH “Community Health Services”) OR (TI “social service*” OR AB “social service*”) OR (TI “social care” OR AB “social care”))

AND

(((MH “United Kingdom+”) OR ((TI “national health service*” OR AB “national health service*”) OR (TI nhs OR AB nhs)) OR ((TI gb OR AB gb) OR (TI “g b” OR AB “g b”) OR (TI britain* OR AB britain*) OR ((TI british* OR AB british*) NOT (TI “british columbia” OR AB “british columbia”)) OR (TI uk OR AB uk) OR (TI “u k” OR AB “u k”) OR (TI “united kingdom*” OR AB “united kingdom*”) OR ((TI england* OR AB england*) NOT (TI “new england” OR AB “new england”)) OR (TI “northern ireland*” OR AB “northern ireland*”) OR (TI “northern irish*” OR AB “northern irish*”) OR (TI scotland* OR AB scotland*) OR (TI scottish* OR AB scottish*) OR (((TI wales OR AB wales) OR (TI “south wales” OR AB “south wales”)) NOT (TI “new south wales” OR AB “new south wales”)) OR (TI welsh* OR AB welsh*)) OR ((TI bath OR AB bath) OR (TI bath’s OR AB bath’s) OR (((TI birmingham OR AB birmingham) NOT (TI alabama* OR AB alabama*)) OR ((TI birmingham’s OR AB birmingham’s) NOT (TI alabama* OR AB alabama*)) OR (TI bradford OR AB bradford) OR (TI bradford’s OR AB bradford’s) OR (TI brighton OR AB brighton) OR (TI brighton’s OR AB brighton’s) OR (TI bristol OR AB bristol) OR (TI bristol’s OR AB bristol’s) OR (TI carlisle* OR AB carlisle*) OR ((TI cambridge OR AB cambridge) NOT ((TI massachusetts* OR AB massachusetts*) OR (TI boston* OR AB boston*) OR (TI harvard* OR AB harvard*))) OR ((TI canterbury OR AB canterbury) NOT (TI zealand* OR AB zealand*)) OR (TI chelmsford OR AB chelmsford) OR (TI chester OR AB chester) OR (TI chester’s OR AB chester’s) OR (TI chichester OR AB chichester) OR (TI coventry OR AB coventry) OR (TI coventry’s OR AB coventry’s) OR (TI derby OR AB derby) OR (TI derby’s OR AB derby’s) OR ((TI durham OR AB durham) NOT ((TI carolina* OR AB carolina*) OR (TI nc OR AB nc))) OR ((TI durham’s OR AB durham’s) NOT ((TI carolina* OR AB carolina*) OR (TI nc OR AB nc))) OR (TI ely OR AB ely) OR (TI ely’s OR AB ely’s) OR (TI exeter OR AB exeter) OR (TI gloucester OR AB gloucester) OR (TI hereford OR AB hereford) OR (TI hull OR AB hull) OR (TI hull’s OR AB hull’s) OR (TI lancaster OR AB lancaster) OR (TI lancaster’s OR AB lancaster’s) OR (TI leeds* OR AB leeds*) OR (TI leicester OR AB leicester) OR ((TI lincoln OR AB lincoln) NOT (TI nebraska* OR AB nebraska*)) OR ((TI lincoln’s OR AB lincoln’s) NOT (TI nebraska* OR AB nebraska*)) OR ((TI liverpool OR AB liverpool) NOT ((TI “new south wales*” OR AB “new south wales*”) OR (TI nsw OR AB nsw))) OR ((TI liverpool’s OR AB liverpool’s) NOT ((TI “new south wales*” OR AB “new south wales*”) OR (TI nsw OR AB nsw))) OR (((TI london OR AB london) NOT ((TI ontario* OR AB ontario*) OR (TI ont OR AB ont) OR (TI toronto* OR AB toronto*))) OR ((TI london’s OR AB london’s) NOT ((TI ontario* OR AB ontario*) OR (TI ont OR AB ont) OR (TI toronto* OR AB toronto*))) OR (TI manchester OR AB manchester) OR (TI manchester’s OR AB manchester’s) OR ((TI newcastle OR AB newcastle) NOT ((TI “new south wales*” OR AB “new south wales*”) OR (TI nsw OR AB nsw))) OR ((TI newcastle’s OR AB newcastle’s) NOT ((TI “new south wales*” OR AB “new south wales*”) OR (TI nsw OR AB nsw))) OR (TI norwich OR AB norwich) OR (TI nottingham OR AB nottingham) OR (TI nottingham’s OR AB nottingham’s) OR (TI oxford OR AB oxford) OR (TI oxford’s OR AB oxford’s) OR (TI peterborough OR AB peterborough) OR (TI plymouth OR AB plymouth) OR (TI plymouth’s OR AB plymouth’s) OR (TI portsmouth OR AB portsmouth) OR (TI preston OR AB preston) OR (TI preston’s OR AB preston’s) OR (TI ripon OR AB ripon) OR (TI salford OR AB salford) OR (TI salisbury OR AB salisbury) OR (TI salisbury’s OR AB salisbury’s) OR (TI sheffield OR AB sheffield) OR (TI sheffield’s OR AB sheffield’s) OR (TI southampton OR AB southampton) OR (TI southampton’s OR AB southampton’s) OR (TI “st albans” OR AB “st albans”) OR (TI stoke OR AB stoke) OR (TI stoke’s OR AB stoke’s) OR (TI sunderland OR AB sunderland) OR (TI sunderland’s OR AB sunderland’s) OR (TI truro OR AB truro) OR (TI wakefield OR AB wakefield) OR (TI wakefield’s OR AB wakefield’s) OR (TI wells OR AB wells) OR (TI westminster OR AB westminster) OR (TI winchester OR AB winchester) OR (TI wolverhampton OR AB wolverhampton) OR ((TI worcester OR AB worcester) NOT ((TI massachusetts* OR AB massachusetts*) OR (TI boston* OR AB boston*) OR (TI harvard* OR AB harvard*))) OR ((TI york OR AB york) NOT ((TI “new york*” OR AB “new york*”) OR (TI ny OR AB ny) OR (TI ontario* OR AB ontario*) OR (TI ont OR AB ont) OR (TI toronto* OR AB toronto*))) OR ((TI york’s OR AB york’s) NOT ((TI “new york*” OR AB “new york*”) OR (TI ny OR AB ny) OR (TI ontario* OR AB ontario*) OR (TI ont OR AB ont) OR (TI toronto* OR AB toronto*)))))) OR ((TI bangor OR AB bangor) OR (TI cardiff OR AB cardiff) OR (TI newport OR AB newport) OR (TI newport’s OR AB newport’s) OR (TI “st asaph” OR AB “st asaph”) OR (TI “st davids” OR AB “st davids”) OR (TI swansea OR AB swansea)) OR ((TI aberdeen OR AB aberdeen) OR (TI dundee OR AB dundee) OR (TI dundee’s OR AB dundee’s) OR (TI edinburgh OR AB edinburgh) OR (TI edinburgh’s OR AB edinburgh’s) OR (TI glasgow OR AB glasgow) OR (TI glasgow’s OR AB glasgow’s) OR (TI inverness OR AB inverness) OR ((TI perth OR AB perth) NOT (TI australia* OR AB australia*)) OR ((TI perth’s OR AB perth’s) NOT (TI australia* OR AB australia*)) OR (TI stirling OR AB stirling) OR (TI stirling’s OR AB stirling’s)) OR ((TI armagh OR AB armagh) OR (TI belfast OR AB belfast) OR (TI lisburn OR AB lisburn) OR (TI londonderry OR AB londonderry) OR (TI derry OR AB derry) OR (TI derry’s OR AB derry’s) OR (TI newry OR AB newry))) NOT ((MH africa+) OR (MH americas+) OR (MH “antarctic regions+”) OR (MH “arctic regions+”) OR (MH asia+) OR (MH oceania+))) NOT ((MH “United Kingdom+”) OR (MH europe))

AND

Limiters - Publication Date: 20090101-20241231

### EconLit (via Proquest)

((TI,AB(predict*) OR TI,AB(determinant*) OR TI,AB(driver*) OR TI,AB(projecti*) OR TI,AB(promot*)) AND (TI,AB(“health services need”) OR TI,AB(“health services demand”) OR TI,AB(need*) OR TI,AB(demand*) OR TI,AB(using) OR TI,AB(utilis*) OR TI,AB(utiliz*) OR TI,AB(access*) OR TI,AB(“health expenditure”) OR TI,AB(resourc*)) AND (TI,AB(“long term care”) OR TI,AB((“social work” OR “social worker” OR “social workers” OR “social works”)) OR TI,AB(“formal care”) OR TI,AB(“informal care”) OR TI,AB(“Home Care Services”) OR TI,AB(“home care”) OR TI,AB(homecare) OR TI,AB(communit*) OR TI,AB(“Community Health Services”) OR TI,AB((“social service” OR “social services”)) OR TI,AB(“social care”)) AND (((TI,AB(“United Kingdom”) OR (TI,AB(“national health service*”) OR TI,AB(nhs)) OR (TI,AB(gb) OR TI,AB(“g b”) OR TI,AB(britain*) OR (TI,AB(british*) NOT TI,AB(“british columbia”)) OR TI,AB(uk) OR TI,AB(“u k”) OR TI,AB((“united kingdom” OR “united kingdoma” OR “united kingdomin” OR “united kingdoms” OR “united kingdomthe”)) OR (TI,AB(england*) NOT TI,AB(“new england”)) OR TI,AB((“northern ireland” OR “northern irelandin” OR “northern irelands” OR “northern irelandthe”)) OR TI,AB((“northern irish” OR “northern irishman”)) OR TI,AB(scotland*) OR TI,AB(scottish*) OR ((TI,AB(wales) OR TI,AB(“south wales”)) NOT TI,AB(“new south wales”)) OR TI,AB(welsh*)) OR (TI,AB(bath) OR TI,AB(bath’s) OR ((TI,AB(birmingham) NOT TI,AB(alabama*)) OR (TI,AB(birmingham’s) NOT TI,AB(alabama*)) OR TI,AB(bradford) OR TI,AB(bradford’s) OR TI,AB(brighton) OR TI,AB(brighton’s) OR TI,AB(bristol) OR TI,AB(bristol’s) OR TI,AB(carlisle*) OR (TI,AB(cambridge) NOT (TI,AB(massachusetts*) OR TI,AB(boston*) OR TI,AB(harvard*))) OR (TI,AB(canterbury) NOT TI,AB(zealand*)) OR TI,AB(chelmsford) OR TI,AB(chester) OR TI,AB(chester’s) OR TI,AB(chichester) OR TI,AB(coventry) OR TI,AB(coventry’s) OR TI,AB(derby) OR TI,AB(derby’s) OR (TI,AB(durham) NOT (TI,AB(carolina*) OR TI,AB(nc))) OR (TI,AB(durham’s) NOT (TI,AB(carolina*) OR TI,AB(nc))) OR TI,AB(ely) OR TI,AB(ely’s) OR TI,AB(exeter) OR TI,AB(gloucester) OR TI,AB(hereford) OR TI,AB(hull) OR TI,AB(hull’s) OR TI,AB(lancaster) OR TI,AB(lancaster’s) OR TI,AB(leeds*) OR TI,AB(leicester) OR (TI,AB(lincoln) NOT TI,AB(nebraska*)) OR (TI,AB(lincoln’s) NOT TI,AB(nebraska*)) OR (TI,AB(liverpool) NOT (TI,AB(“new south wales*”) OR TI,AB(nsw))) OR (TI,AB(liverpool’s) NOT (TI,AB(“new south wales*”) OR TI,AB(nsw))) OR ((TI,AB(london) NOT (TI,AB(ontario*) OR TI,AB(ont) OR TI,AB(toronto*))) OR (TI,AB(london’s) NOT (TI,AB(ontario*) OR TI,AB(ont) OR TI,AB(toronto*))) OR TI,AB(manchester) OR TI,AB(manchester’s) OR (TI,AB(newcastle) NOT (TI,AB(“new south wales*”) OR TI,AB(nsw))) OR (TI,AB(newcastle’s) NOT (TI,AB(“new south wales*”) OR TI,AB(nsw))) OR TI,AB(norwich) OR TI,AB(nottingham) OR TI,AB(nottingham’s) OR TI,AB(oxford) OR TI,AB(oxford’s) OR TI,AB(peterborough) OR TI,AB(plymouth) OR TI,AB(plymouth’s) OR TI,AB(portsmouth) OR TI,AB(preston) OR TI,AB(preston’s) OR TI,AB(ripon) OR TI,AB(salford) OR TI,AB(salisbury) OR TI,AB(salisbury’s) OR TI,AB(sheffield) OR TI,AB(sheffield’s) OR TI,AB(southampton) OR TI,AB(southampton’s) OR TI,AB(“st albans”) OR TI,AB(stoke) OR TI,AB(stoke’s) OR TI,AB(sunderland) OR TI,AB(sunderland’s) OR TI,AB(truro) OR TI,AB(wakefield) OR TI,AB(wakefield’s) OR TI,AB(wells) OR TI,AB(westminster) OR TI,AB(winchester) OR TI,AB(wolverhampton) OR (TI,AB(worcester) NOT (TI,AB(massachusetts*) OR TI,AB(boston*) OR TI,AB(harvard*))) OR (TI,AB(york) NOT (TI,AB((“new york” OR “new yorker” OR “new yorkers” OR “new yorks” OR “new yorkshire” OR “new yorkthe” OR “new yorkthis”)) OR TI,AB(ny) OR TI,AB(ontario*) OR TI,AB(ont) OR TI,AB(toronto*))) OR (TI,AB(york’s) NOT (TI,AB((“new york” OR “new yorker” OR “new yorkers” OR “new yorks” OR “new yorkshire” OR “new yorkthe” OR “new yorkthis”)) OR TI,AB(ny) OR TI,AB(ontario*) OR TI,AB(ont) OR TI,AB(toronto*)))))) OR (TI,AB(bangor) OR TI,AB(cardiff) OR TI,AB(newport) OR TI,AB(newport’s) OR TI,AB(“st asaph”) OR TI,AB(“st davids”) OR TI,AB(swansea)) OR (TI,AB(aberdeen) OR TI,AB(dundee) OR TI,AB(dundee’s) OR TI,AB(edinburgh) OR TI,AB(edinburgh’s) OR TI,AB(glasgow) OR TI,AB(glasgow’s) OR TI,AB(inverness) OR (TI,AB(perth) NOT TI,AB(australia*)) OR (TI,AB(perth’s) NOT TI,AB(australia*)) OR TI,AB(stirling) OR TI,AB(stirling’s)) OR (TI,AB(armagh) OR TI,AB(belfast) OR TI,AB(lisburn) OR TI,AB(londonderry) OR TI,AB(derry) OR TI,AB(derry’s) OR TI,AB(newry))) NOT (TI,AB (africa) OR TI,AB(americas) OR TI,AB(“antarctic regions”) OR TI,AB(“arctic regions”) OR TI,AB(asia) OR TI,AB(oceania))) NOT (TI,AB(“United Kingdom”) OR TI,AB(europe)))) AND pd(20090101-20241231)

### Applied Social Science Index and Abstracts, ASSIA (via ProQuest)

((TI,AB(predict*) OR TI,AB(driver*) OR TI,AB(determinant*) OR TI,AB(projecti*) OR TI,AB(promot*)) AND (MESH.EXACT(“Health Services Needs and Demand”) OR TI,AB(need*) OR TI,AB(demand*) OR TI,AB(using) OR TI,AB(utilis*) OR TI,AB(utiliz*) OR TI,AB(access*) OR MESH.EXACT(“Health Expenditures”) OR TI,AB(resourc*)) AND (MESH.EXACT(“long term care”) OR TI,AB(“long term care”) OR MESH.EXACT(“Social Work”) OR TI,AB((“social work” OR “social worker” OR “social workers” OR “social works”)) OR TI,AB(“formal care”) OR TI,AB(“informal care”) OR MESH.EXACT(“Home Care Services”) OR TI,AB(“home care”) OR TI,AB(homecare) OR TI,AB(communit*) OR MESH.EXACT(“Community Health Services”) OR TI,AB((“social service” OR “social services”)) OR TI,AB(“social care”))) AND (location.exact(“United Kingdom--UK” OR “England” OR “Wales” OR “Scotland” OR “Northern Ireland”) AND pd(20090101-20241231))

### Campbell Collaboration

#### Search 1

Campbell Subject Category: Ageing AND Systematic reviews AND publication date: 2009 – 2024

#### Search 2

Campbell Subject Category: Social welfare AND Updated systematic reviews AND publication date: 2009 – 2024

#### Search 3

Campbell Subject Category: Social welfare AND Systematic reviews AND publication date: 2009 – 2024

